# Clinical Presentation, Complication and Outcome of Black Stone (Paraphenylene Diamine) Poisoning

**DOI:** 10.1101/2020.10.24.20218800

**Authors:** Hakim Ali Abro, Sultan Ahmed Chandio, Azizullah Jalbani, Sheeraz Ali Buriro, Aafia Saeed, Faizan Shaukat

## Abstract

**Introduction:** In recent time, developing countries of South Asia and Africa have seen significant increase in ingestion of Para-Phenylene Diamine (PPD), locally known as Kala Pathar, either accidental or for suicide. Through this study, we aim to study the clinical presentations and outcomes among patients who have ingested PPD.

**Method:** This retrospective case series study was conducted in a tertiary care hospital of Pakistan, from April 2013 to August 2017. Data of patients of PPD poisoning was archived from the hospital’s medical records. Around 174 consecutive cases were included in the study. Patients were evaluated based on self-administrated proforma.

**Result:** Out of 174 cases of PPD poisoning that were identified, 57(32.8%) were males and 117(67.2%) were females. The mean age ± SD (range) of the patients was 24.16±9 (10 to 70) years. Approximately 170 (97.8%) patients used PPD for suicidal intention. The most common presentation was facial swelling which was present in 144(82.8%) patients followed by dysphagia in 143(82.2%) patients. Complications include metabolic acidosis in 50 (28.7%) patients and aspiration pneumonia in 36 (20.7%) patients. A total of 101 (58%) improved, while others were either referred or left against medical advice (LAMA).

**Conclusion:** Increasing incidence of ingestion of PPD for suicide warrants the regulatory authorities to restrict the use of PPD in hair dyes and implement strict measures to educate masses and curtail the easy access of such poisonous substances among common people.

## Introduction

Globally, around one million deaths occur every year due to self-murder and in last half century the trend of suicide is escalated by 60% [1, 2]. Different methods of committing self-harm are being practiced in different societies and many majorities of the people choose suicide for ending their life. Thus, currently numerous health care facilities are facing substantial number of poisoning cases in their emergencies [3]. Different trends of committing suicide are prevailing around the universe and mostly choice depends upon the social status as pesticides are commonly ingested in eastern and lower socio-economic class while in high socio-economic and influential societies, opiates and tranquillizers are used for this purpose [4, 5]. South Asia and Africa are witnessing a increase number of people committing or attempting suicide with Paraphenylene diamine (PPD) which is responsible for enormous number of the deaths in these regions [6-8].

PPD is an organic by product of aniline dye with formula of C_6_H_4_(NH_2_)_2_ and when oxidized in air, its color changes into black from white [9]. Principally, it enhances the dyeing process and is an ingredient of many hair color products currently available in market. Historically, PPD was mixed with henna for coloring hair and when ingested, it can prove to be lethal [10]. In developing countries, common man has easy access to get PPD from market for suicidal intention with no check and balance on its trade [11]. PPD is active constituent of Kala Pathar (Meaning Black Stone in Urdu) and when taken orally or used locally, it has harmful effects [10]. The active components of kala pathar includes 4% PPD, resorcinol, propylene glycol, ethylene-diamine-tetraacetic acid (EDTA), sodium, liquid paraffin, cetostearyl alcohol, sodium lauryl sulphate, herbal extracts, preservatives, and perfumes [12]. Out of these, few have notorious fatal effects while poisonous profile of other substances is still in question. PPD principally produces its damaging effects on all the major organs of body like kidneys, liver, heart, lungs, muscles, and mucous membrane. Individuals affected with PPD poisoning rapidly develop edema of oral cavity and upper respiratory tract by producing local effects followed by rhabdomyolysis and acute kidney insult due to precipitation of its toxic breakdown products in renal tubules [13, 14]. Some of the studies have shown that persons using commercially available hair color have increased likehood to develop multiple myeloma, non-Hodgkin’s Lymphoma, acute leukemia, and bladder cancer [15].

Currently, there is limited data available on this poisoning as it has emerged as a new trend in our local hospitals. So, the rationale of this study is to evaluate the clinical presentation and outcome of the PPD poisoning in our part so that effective preventive and treatment strategies can be implemented at the onset which would certainly decrease the morbidity and mortality associated with this deadly poisoning. This study can lay down a base for local administrative authorities to consider formulating a law for strict handling of the PPD poisoning at local level and for education of the population.

## Material and Methods

This retrospective case series study was conducted at the medical intensive care unit of Medical Unit-I of Chandka Medical College Hospital, Pakistan. All the PPD poisoning patients with both gender and more than 10 years age admitted to medical ICU were included. The diagnosis of PPD poisoning was based on clinical findings and information taken from the patient’s family and friends. Patients with history of liver, cardiac and renal disease were excluded from study.

After approval by ethical review committee of the hospital, patient’s data was retrieved from their medical record files. A pro-forma was used to collect data including demographic features (age, sex, marital status, socio economic status), clinical features including examination findings (especially cervico-facial edema, dysphagia, respiratory difficulty, syncope/coma, fits, color of urine, blood pressure, anemia, edema, oliguria), laboratory findings (complete blood count, liver function test, creatinine kinase [CK], lactate dehydrogenase [LDH], glucose, urea, creatinine, electrolytes and electrocardiogram [ECG]), mode of intoxication (accidental or suicide) and route of intoxication (gastrointestinal system, skin) was recorded. Hospitalization time after the ingestion, intent of ingestion (accidental or intentional) total hospital stays, amount of poison ingested, complications and final outcomes in the form of improvement, referral and death were also recorded.

Data was analyzed on computer using SPSS software version 19. The detailed socio-demographic data like age, gender, marital status and other clinical data like duration, route and intent of poisoning was recorded. The mean and standard deviation (SD) was calculated for age and duration of poisoning. Frequencies and percentages were calculated for gender, clinical presentation including examination findings, route, intent of poisoning and outcome in the form of improvement and discharge from hospital, referral and death was be calculated. Post stratification Chi square test was be used to see for significance and p value of ≤ 0.05 meant null hypothesis is not applicable and there is a difference between variables.

## Results

Total 174 consecutive cases with PPD poisoning were observed during the study period. Out of them 57(32.8%) were males and 117(67.2%) were females. The mean age ± SD (range) of the patients was 24 ±9 (10 to 70) years with range of minimum 10 years and maximum 70 years and majority of the patients 82 (47.1%) were in the age group 21-30 years followed by 66(37.9%) patients were in the age group 21 to 30 years, 17(9.8%) were seen in age group 31 to 40 years, 7(74.0%) were observed in the age group 41 to 50 years and only 1(0.6%) patient was seen tin the group 51 to 60 years or above > 60 years.

Regarding the marital status 106 (60.9%) were unmarried while 68 (39.1%) were married. As far as socioeconomic status of the cases is concerned, 137 (78.7%) belonged to poor background, 35 (20.1%) were middle class cases and 2(1.1%) belonged to high class.

Regarding reasons of ingestion, suicidal intention was observed in 170 (97.8%), in 2 (1.1%) patients it was accidental and homicidal, respectively. All 174 patients ingested local made, black stone-based hair dye via oral route.

The time interval to reach hospital ranged from 1 to 24 h with a mean duration of 5.36 ± 4.67 hours. Out of 174 patients, 138 (79.3%) were brought to hospital emergency less than 6 hours, 33(19.0%) patients were brought between 6 to 18 hours and only 3 (1.7%) patients reached the hospital after more than 18 hours (table 1)

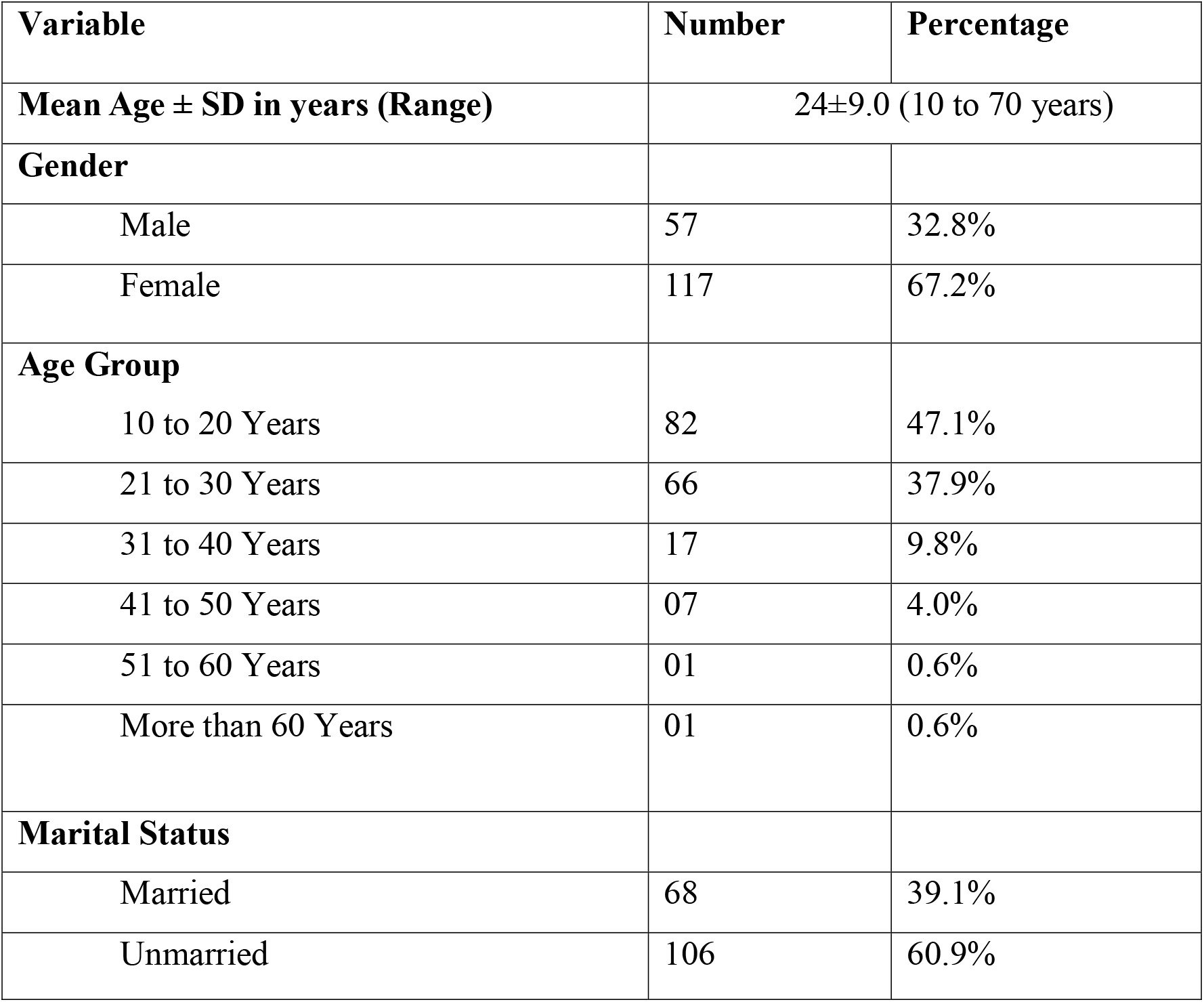
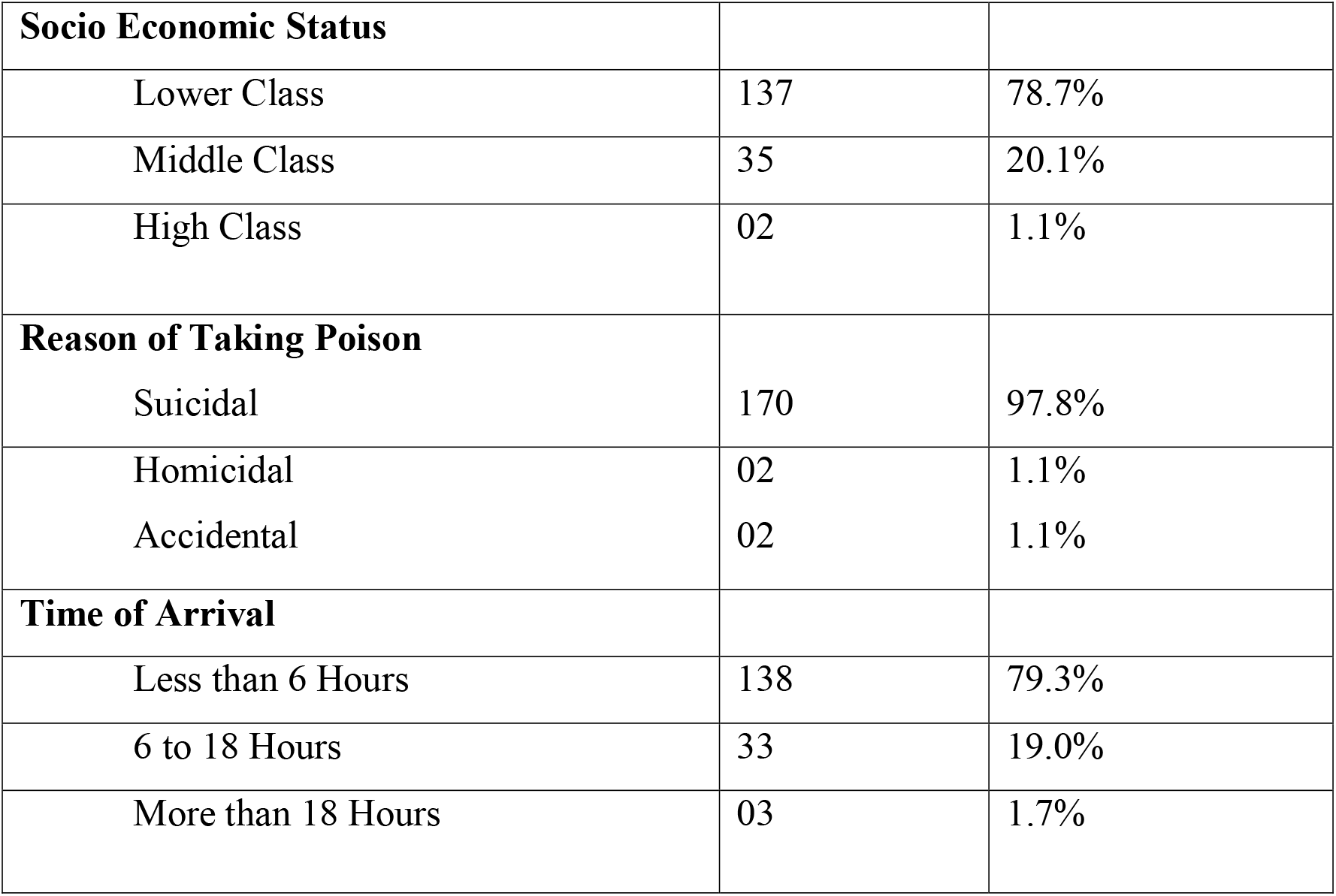

The clinical presentation of patients was proportionately associated with the amount and the type of dye consumed. Most common presentation was facial swelling which was present in 144(82.8%) patients and dysphagia was second most common symptom which was seen in 143(82.2%) patients. Respiratory difficulty was present in 142(81.6%) patients. Other common presentations were tachycardia, pain and rigidity of limb, cola brown color urine, palpitation, decrease urine output, nasal twang of voice, oliguria, pre-syncope, chest Pain, anuria, nasal regurgitation, and convulsion. These patients had a history of immediate swallowing of large amount of hair dye and developed features suggestive of myocarditis (Table 2).

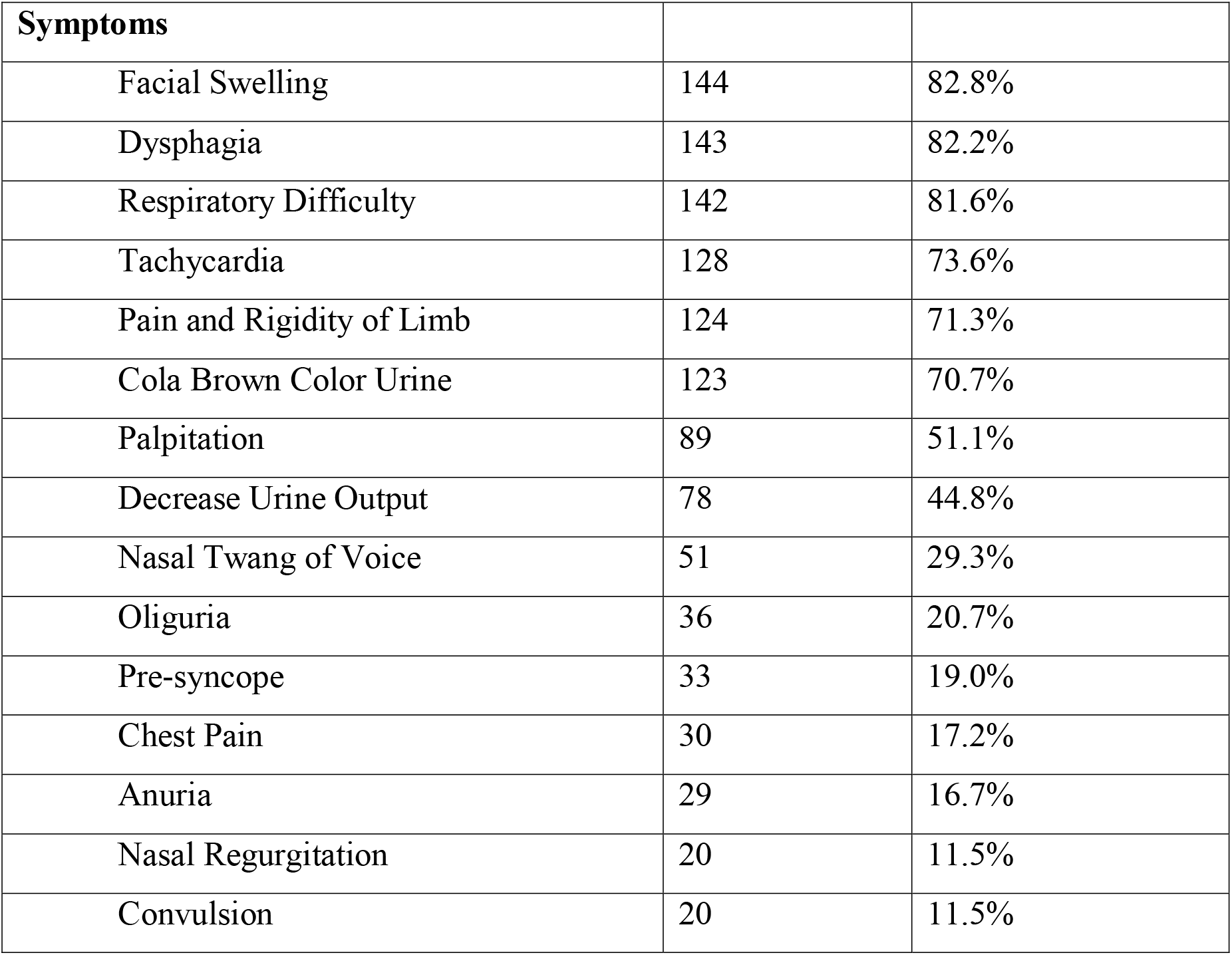

With regards to clinical examination, out of 174 patients, mean temperature ± standard deviation (SD) was 98.8±0.7, mean pulse ± SD was 110±28, mean respiratory rate ± SD was 26±07, mean blood pressure (BP) systolic ± SD was 131±27 and mean BP diastolic ± SD was 80.3±19. Facial edema was seen in 144(82.8%) cases followed by dehydration in 88(50.6%) cases, cyanosis in 25(14.4%), jaundice in 22(12.6%) and anemia was seen in 9(5.2%) cases (table 3).

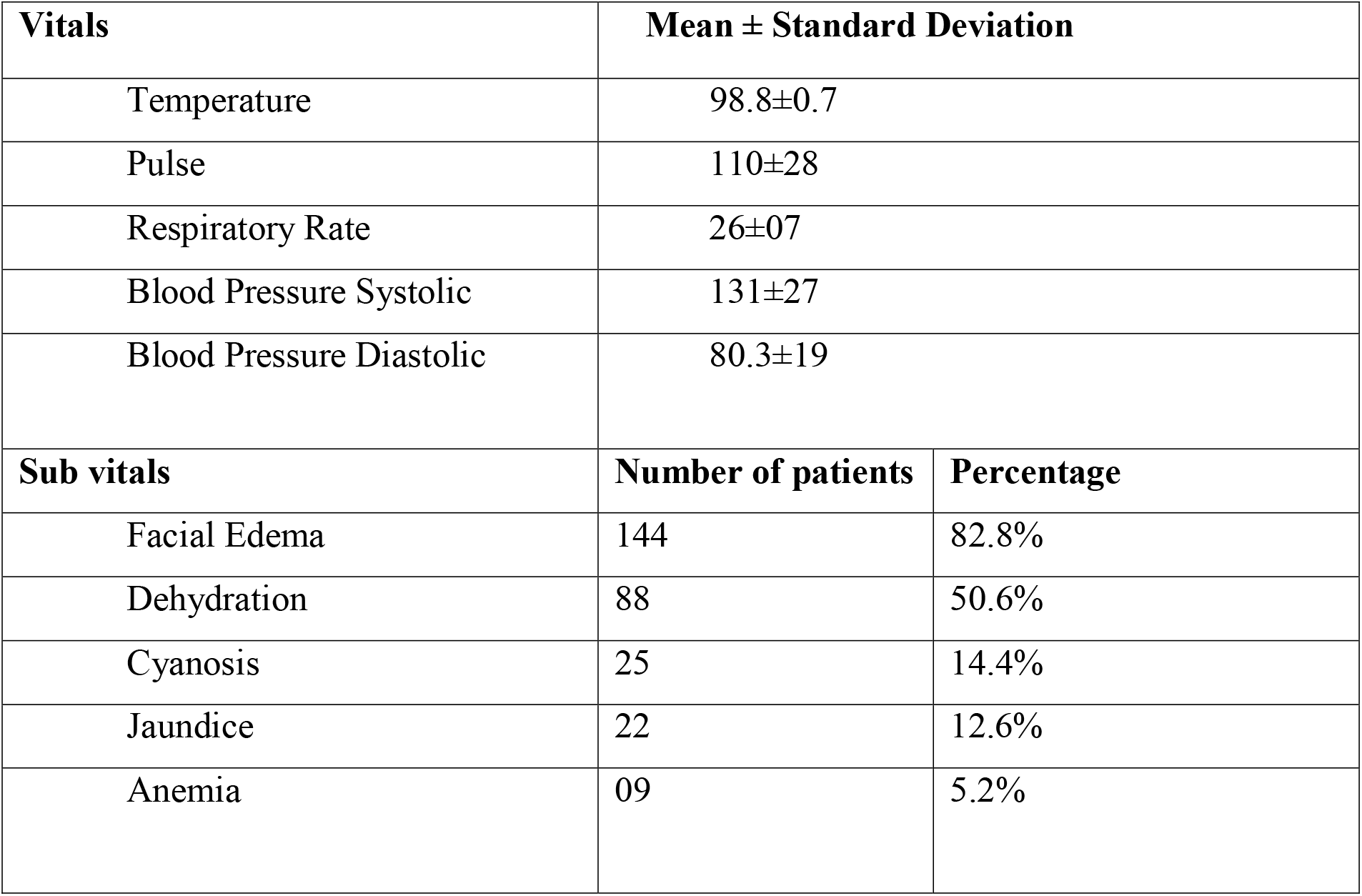

Regarding laboratory investigations, the mean ± standard deviation of complete blood count (CBC) such as hemoglobin (%), total leukocyte count (TLC), serum platelet levels were 12.6±2.4, 18376±12356 and 287125±100303 respectively. Mean random blood sugar level was 166.0±78.0 and LFTs (liver function tests) such as total bilirubin (mg/dL), Alanine Amino-tranferase (U/L), Alkaline Phosphatase (U/L) levels were 0.8±0.5, 144±166, and 162±70 respectively. Renal function was estimated as mean ± SD of serum urea (mg/dL) and creatinine (g/dL) levels, which were 42.6±32 and 1.3±1.0.

Creatine phosphokinase (CPK) levels were found to be increased, i.e. above 300 IU. Mean CPK ± SD (range) was 73591±127371 (3200 to 1055010). CPK levels gradually decline reaching normal levels within an average period of 10 days. Chest X-ray showed features of aspiration in 6(3.5%) patients and normal chest X-ray was seen in 149(85.6%) patients. ECG abnormalities like sinus tachy-arrhythmias were observed in 14(8.0%) patients and brady-arrhythmia in 9(5.2%) patients.

In this study, 11 of 174 patients (6.3%) developed acute hepatitis within 6 hours of poison intake. Metabolic acidosis was evident in 50 of 174 cases (28.7%). Around 36(20.7%) of 174 patients with CPK levels more than 10,000 IU/L developed renal failure (elevation of serum creatinine) and required dialysis, while 36(20.7%) patients developed aspiration pneumonia. Arrhythmia was seen in 30(17.2%) patients. Hypocalcaemia was observed in 12(6.9%) patients. Only 1(0.6%) patient suffered from bleeding (table 4).

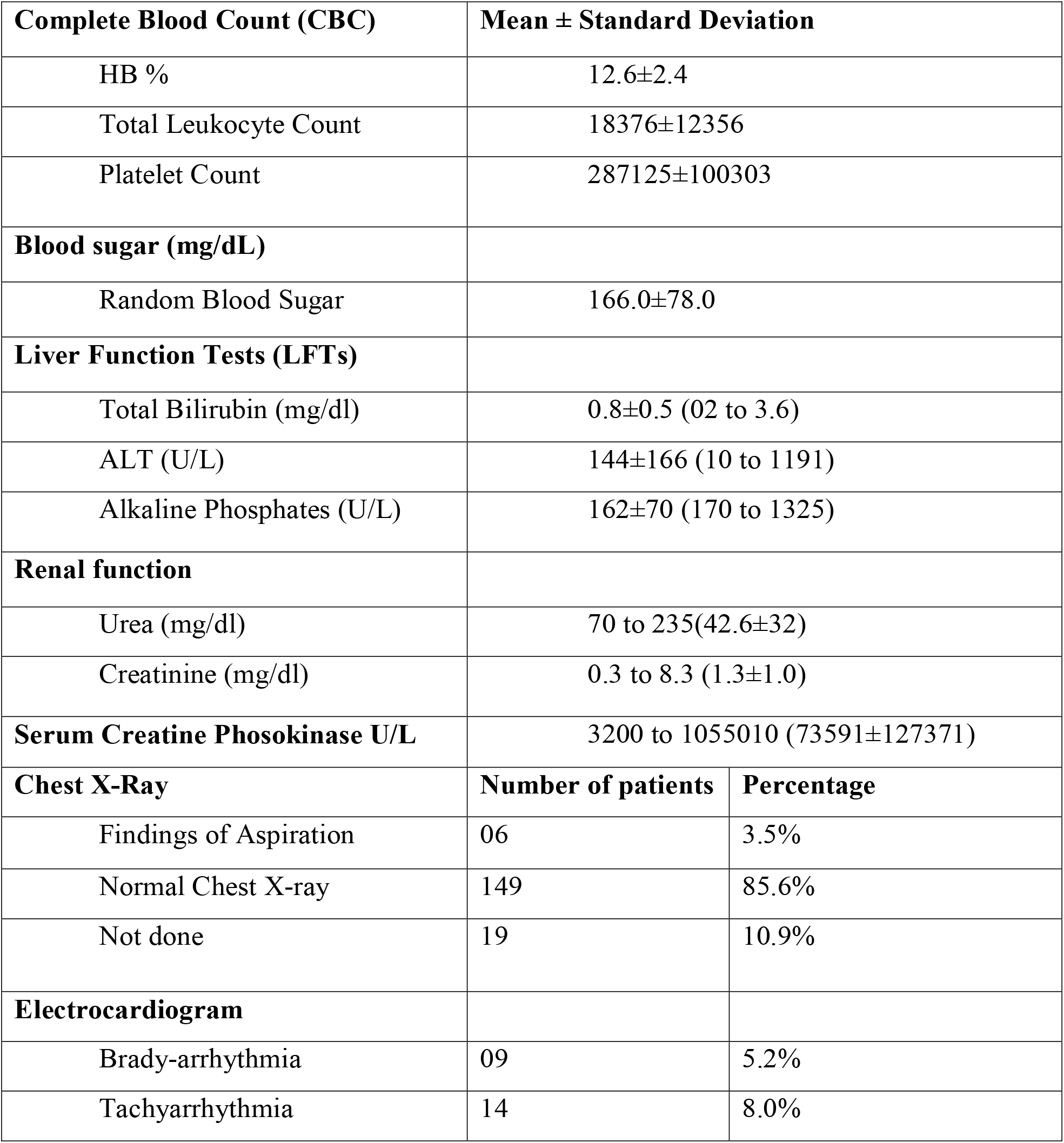
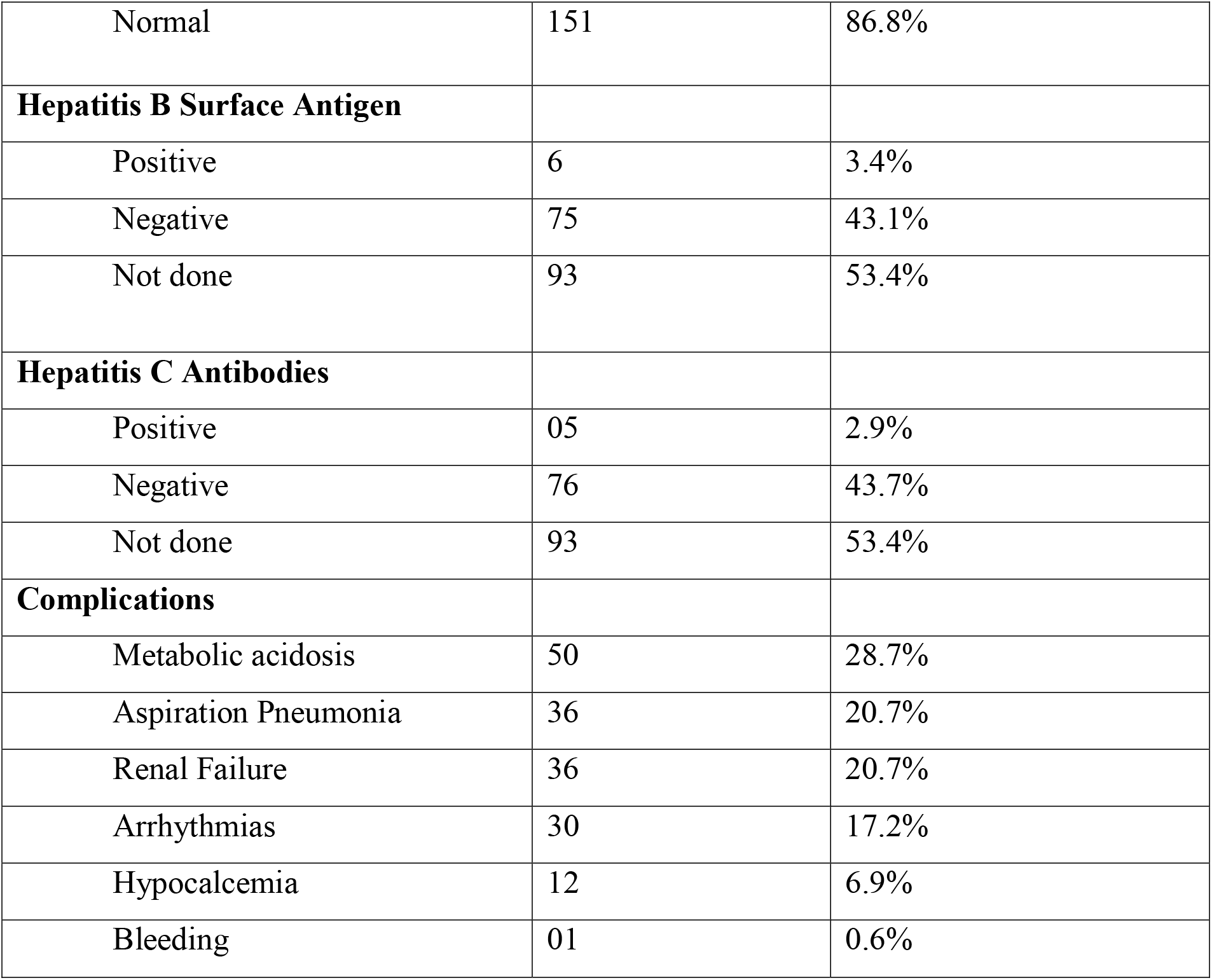

| **Complete Blood Count (CBC)** |  | **Mean ± Standard Deviation** |
| --- | --- | --- |
| HB % |  | 12.6±2.4 |
| Total Leukocyte Count |  | 18376±12356 |
| Platelet Count |  | 287125±100303 |
| **Blood sugar (mg/dL)** |  |  |
| Random Blood Sugar |  | 166.0±78.0 |
| **Liver Function Tests (LFTs)** |  |  |
| Total Bilirubin (mg/dl) |  | 0.8±0.5 (02 to 3.6) |
| ALT (U/L) |  | 144±166 (10 to 1191) |
| Alkaline Phosphates (U/L) |  | 162±70 (170 to 1325) |
| **Renal function** |  |  |
| Urea (mg/dl) |  | 70 to 235(42.6±32) |
| Creatinine (mg/dl) |  | 0.3 to 8.3 (1.3±1.0) |
| **Serum Creatine Phosokinase U/L** |  | 3200 to 1055010 (73591±127371) |
| **Chest X-Ray** | **Number of patients** | **Percentage** |
| Findings of Aspiration | 06 | 3.5% |
| Normal Chest X-ray | 149 | 85.6% |
| Not done | 19 | 10.9% |
| **Electrocardiogram** |  |  |
| Brady-arrhythmia | 09 | 5.2% |
| Tachyarrhythmia | 14 | 8.0% |
| Normal | 151 | 86.8% |
| **Hepatitis B Surface Antigen** |  |  |
| Positive | 6 | 3.4% |
| Negative | 75 | 43.1% |
| Not done | 93 | 53.4% |
| **Hepatitis C Antibodies** |  |  |
| Positive | 05 | 2.9% |
| Negative | 76 | 43.7% |
| Not done | 93 | 53.4% |
| **Complications** |  |  |
| Metabolic acidosis | 50 | 28.7% |
| Aspiration Pneumonia | 36 | 20.7% |
| Renal Failure | 36 | 20.7% |
| Arrhythmias | 30 | 17.2% |
| Hypocalcemia | 12 | 6.9% |
| Bleeding | 01 | 0.6% |

In our study, most of the patients i.e. 128(73.6%) patients had duration of hospital stay less than 7 days, 37(21.2%) patients had duration of hospital stay 7 to 14 days and 9(5.2%) patients had hospital stay more than 14 days (Table 5).

|  |  |  |
| --- | --- | --- |
| **Duration of Hospital Stay** |  |  |
| Less than 7 days | 128 | 73.6% |
| 7 to 14 days | 37 | 21.2% |
| More than 14 days | 9 | 5.2% |

Overall, mortality was noted in 52 (29.9%); out of these, 44 patients had hospital stay less than 7 days, 4 patients had duration of hospital stay from 7 to 14 days and more than 14 days respectively. Around 101 (58.0%) patients achieved complete clinical recovery. Remaining 21 patients were referred or left against medical advice (LAMA) (table 6).

### Hospital Stay to Outcome

| Character | Improved | Expired | Referred | Lama | Total |
| --- | --- | --- | --- | --- | --- |
| Less than 7 days | 67(52.3%) | 44(34.3%) | 08(6.3%) | 09(7.1%) | 128(100%) |
| 7 to 14 days | 30 (81.0%) | 04(10.8%) | 02(5.5%) | 01(2.7%) | 37(100%) |
| More than 14 days | 04(44.4%) | 04(44.4%) | 01(11.2%) | 0(0%) | 09(100%) |
| **Total** | **101 (58%)** | **52 (29.9%)** | **11(6.4%)** | **10(5.7%)** | **174(100%)** |

## Discussion

Kala Pathar (PPD) poisoning is emerging means of intentional self-harm with high mortality in Pakistan [16]. Easy availability, low cost, and harmful effects of this hair dye ingestion makes it a frequent choice for committing self-harm [16]. It is used to commit suicide or can be ingested accidentally. It can also be mixed in edibles with criminal intentions and given to innocent victims.

In the present study, a sample of 174 poisoned patients by PPD (hair dye) was studied. We found that majority of the patients 82 (47.1%) were in the age group 21-30 years. Elgamel AA et al. [17] reported a peak incidence at 17-27 years, while Musa et al. [18] reported the peak incidence between 21 and 30 years. This may be due to stress which increases at this age. In comparison to study conducted by Sakuntala et al [19], who reported same observation in his study that prevalence was more in the age group of 21– 30 years with female preponderance of 80.64%. According to Nisar Khan et al, young age-group (mean 22.08±6.42 year) is the main sufferer of kala-pathar poisoning [20]. It is consistent with many other studies. Akbar et al [8] reported the mean age as 25.5±4.56 years, Chrispal et al [21] as 27.75 years, and Suliman et al [22] as 40 years. This finding is also in accordance with the WHO report that young age group is more vulnerable to have self-harm in the low- and middle-income countries [20].

Most patients in this study were young females who ingested the poison with the intent of suicide or self-harm. Elgamel AA et al. also found that that (80.5%) were females and the (19.5%) were males [17]. Nisar Khan et al. documented that the female gender was primarily affected by this poisoning with male to female ratio of 1:18 [20]. A study from Sudan by Suliman et al. reported male to female ratio as 1:4 [22]. A study from Hyderabad, India by Sakuntala et al. reported it in females 80.64% as compared to males 18.75% [19]. A study from Multan, Pakistan by Akbar et al also showed similar results [8]. All the five patients in their case-series were females [8]. The explanation for female preponderance could be the use of kala-pathar as a low cost and easily available hair dye. Besides this, females are more exposed to gender inequities and social pressures in the developing countries. In the present study, 106 (60.9%) were unmarried, while 68 (39.1%) were married. Nisar Khan et al. showed, 71.1% patients were unmarried [20].

In our study, we found that suicidal intention was observed in 170 (97.8%) patients, while in 2 (1.1%) patients it was accidental and homicidal each. Similarly, Nisar Khan et al. in his study revealed a high proportion of PPD intoxication (94.74%) based on suicidal intention as well, which favors the contribution of social factors toward this event [20]. This finding is also consistent with other studies i.e. Akbar et al [8] from Pakistan identified suicidal intention in 60% cases, and Suliman et al [22] from Sudan identified suicidal ideation in 84% cases. This shows that PPD as accidental intoxicant is not common in the developing world. Khaskheli MS et al. reported that the intent of poisoning was suicidal in 98.94% of his cases; however psychological evaluation was found to be normal in all these patients [23]. This indicated that most of suicidal attempts were impulsive precipitated by either scolding from parents, family quarrels or socioeconomic reasons [24]. In our study, all patients were exposed to hair dye (black stone) through oral route probably due to ease of administration.

In the current study, facial swelling edema was the most frequent which required emergency tracheostomy. Dysphagia (82.2%), respiratory difficulty (81.6%) and tachycardia (73.6%) were among other common complaints. A prospective study by Jain et al, comprising of 1020 patients with hair dye poisoning in India, reported that the development of severe facial edema occurred in 73.03%, dark urine in 53.82%, and muscular pain in 47.05% patients [25]. Moreover, facial edema was the first symptom to develop as observed in other studies as well [21, 26]. Suliman et al. reported acute renal failure in 60% of their patients [22]. Shock was another important feature due to PPD poisoning which occurred in 18.5% patients in our series.

The mean time of arrival at the hospital in the present study was 5.36 ± 4.67 hours. This is comparable to the mean arrival time of 8.9 ±10.9 hours stated in a study in India [27]. Nisar Khan et al [20] showed the mean time to reach the hospital was 4.68±5.31 hours.

In this study, the overall mortality was noted in 52 (29.9%) patients; out of these, 44 patients had hospital stay less than 7 days, 4 patients had duration of hospital stay from 7 to 14 days and more than 14 days respectively. Approximately 101 (58.0%) patients achieved complete clinical recovery. It was comparable to Kallel et al. and Sakuntala et al [19, 26].

PPD poisoning is more pronounced among youngsters, illiterate and poor people of the developing countries especially in rural areas. The high rate of morbidity and mortality has raised health concerns associated with PPD poisoning. Intensive supportive care and appropriate interventions including tracheostomy is the mainstay of management. PPD containing hair dyes are a great hazard and have been banned in countries like Germany, France and Sweden. However, in Pakistan it is still commonly used due to easy availability and access in many parts and needs to be banned.

## Data Availability

Data is available on request

